# Electrocardiographic safety of daily Hydroxychloroquine 400mg plus Azithromycin 250mg as an ambulatory treatment for COVID-19 patients in Cameroon

**DOI:** 10.1101/2020.06.24.20139386

**Authors:** Liliane Mfeukeu-Kuate, William Ngatchou, Mazou Ngou Temgoua, Charles Kouanfack, Daniel Lemogoum, Joel Noutakdie Tochie, Armel Zemsi, Lauriane Fomete, Skinner Lekelem, Sylvain Zemsi, Joelle Sobngwi, Thierry Ntandzi, Christian Ngongang Ouankou, Yves Wasnyo, Antoinette Ntsama Assiga, Jan René Nkeck, Ahmadou Musa Jingi, Magellan Guewo, Eric Walter Pefura Yone, Charlotte Moussi Omgba, Paul Owono Etoundi, Jean Cyr Yombi, Alain Menanga, Samuel Kingue, Jacqueline Ze Minkande, Jean Claude Mbanya, Pierre Ongolo Zogo, Pierre Joseph Fouda, Eugène Sobngwi

**Author notes:** **Corresponding author:** Professor Eugene Sobngwi, Faculty of Medicine and Biomedical Sciences, University of Yaounde 1, and Yaounde Central Hospital, Cameroon.

## Abstract

**Objective:** To determine the early electrocardiographic changes in a cohort of ambulatory cameroonian COVID-19 patients treated with hydroxychloroquine and Azithromycin.

**Design:** Prospective study.

**Setting:** Treatment centres of the city of Yaounde, Cameroon, from May 7th to 24th 2020.

**Participants:** We enrolled 51 consecutive confirmed COVID-19 on RT-PCR who having mild forms of COVID-19 and treated by hydroxychloroquine 200mg twice daily during seven days plus Azithromycin 500 mg the first day and 250 mg the remaining 4 days as per national standard.

**Main outcomes measures:** The primary end-point was the change in QTc interval between day 0 (D0), day 3 (D3) and day 7 (D7). Secondary endpoints were changes in all other cardiac electrical conductivity patterns and the occurrence of clinical arrhythmic events during the course of treatment.

**Results:** The population (29 men and 22 women) was aged 39 ± 11 years (range 17 to 61 years). Mean Tisdale score was 3.35±0.48. No significant change from baseline (D0) of QTc was observed at D7 (429±27 ms at D0 versus 396±26 ms at D7; p=0.27). A reduction of heart rate was observed between the D0 and D7 (75±13 bpm versus 70±13 bpm, p = 0.02) with increased QRS duration between D0 and D7 (95±10 ms versus 102±17 ms, p = 0.004). No symptomatic arrhythmic events occurred during the treatment course.

**Conclusions:** No life-threatening modifications of the QT interval was observed in non-severe COVID-19 patients treated ambulatory with hydroxychloroquine and azithromycin. Studies are needed in critical-ill and older patients.

## Introduction

Coronavirus disease 19 (COVID-19) is a global public health emergency. Since March 2020, it is recognized as a pandemic.^1^ By May 23, 2020, the total number of COVID-19 cases was 5 067 579 in the world with 332 711 global deaths.^2^ The disease can range from mild to severe and critical forms. It has been reported that Severe Acute Respiratory Coronavirus 2 (SARS-CoV 2), the viral pathogen of COVID-19 affects multiple organs and that one of the major causes of death attributable to COVID-19 was cardiovascular events.^3,4,5^ The interaction between COVID-19 and the cardiovascular system has been attributed to both the direct effect of SARS-CoV2 in endocardial and myocardial cells through Angiotensin Converting Enzyme 2 (ACE2), but also via an indirect inflammatory mechanism with massive cytokine release. To date, there is no effective treatment for COVID-19. Treatment regimens such as the association of hydroxychloroquine and azithromycin are extensively used in Africa despite controversies on the efficacity and cardiovascular safety.^6,7^ In this preliminary report, we aimed to investigate short term electrocardiographic safety of the combination of 400 mg Hydroxychloroquine and Azithromycin among a population with high likelihood of previous exposure to chloroquine as part of malaria control efforts.

## Methods

### Study design, setting, study duration, population

To inform decision about the discontinuation or not of the Hydroxychloroquine and Azithromycine based COVID-19 treatment protocol, we conducted a prospective study in approved COVID-19 treatment centres in the city of Yaounde, Cameroon. From May 7th through 24th 2020, all consenting consecutive consenting confirmed COVID-19 patients (by RT-PCR on nasopharyngeal swab) on ambulatory treatment due to the non-severe form of the disease were enrolled and follow-up for 7 days. The national standard for non-severe COVID-19 includes oral hydroxychloroquine 200mg twice daily for seven days and Azithromycin 500 mg the first day and 250 mg from day 2 through 5.

After an interviewer-administered health status questionnaire and clinical examination, a trained nurse performed a 12-leads electrocardiography (ECG) prior to treatment (D0). We excluded all patients with a baseline QTc > 450 msec, a Tisdale score > 10, the use of concomitant drug that prolong the QTc interval or use of Hydroxychloroquine or Azithromycin within 10 days prior to inclusion. Subsequent ECG recording was performed on Day 3 and Day 7 after starting treatment with Hydroxychloroquine and Azithromycin. The analysis of ECG was done by three independent consultant cardiologists (LMK,WN,MNT), and findings communicated to participants. A treatment discontinuation was planned in case of prolongation of QTc interval > 500ms or in case of major clinical arrhythmic events.

### Variables Assessed

Study variables included sociodemographic (age and gender), clinical (blood pressure, heart rate and oxygen saturation) characteristics, and ECG findings such as the heart rhythm, heart rate, PR-interval, QRS duration and electric axis, QT-interval corrected by Bazett formula (QTc),^8^ the ST-segment and the Tisdale arrhythmia risk score.^9^

### Primary endpoint

change in QTc-interval from baseline (D0) at D3 and D7.

### Secondary endpoints

changes of all other electrical patterns of the ECG and the occurrence of clinical arrhythmic events during the treatment course.

### Statistical analysis

Fisher test was used to compare changes in categorical variables and Mann-withney tests for quantitative variables. Linear regression model was used to assess change between D0, D3 and D7 with p-value < 0.05 considered significant.

### Ethics

The study protocol received clearance from the National Ethics Committee on May 6^th^ prior to inclusion of patients and all participants were adults who gave written informed consent before enrolment.

## Results

### General characteristics

A total of 51 participants were enrolled. The mean age of the participants was 39 ± 11 years (ranging from 17 to 61 years). The male gender was predominant (n=29, 57%). Participants had few co-morbidities including hypertension in 5.9%. Fever and cough were the most frequently reported symptoms with 21.6% and 23.5% respectively (Table 1). Mean blood pressure was 130±25 (systolic) / 84±19 (diastolic) mmHg, and mean PaO2 at enrolment 98±1%. The risk of arrhythmia was low as evidenced by a mean Tisdale score of 3.35±0.48. At baseline, electrocardiographic recordings presented few abnormalities with left ventricular hypertrophy in 9.8% and the depression of ST-segment in 2 patients.

**Table 1:** General characteristics of the participants.

| **Variables** | **Frequency**<br>**N= 51** | **Percentage**<br>**(%)** |
| --- | --- | --- |
| **Level of Education** |  |  |
| Primary | 5 | 10 |
| Secondary | 12 | 24 |
| University | 33 | 66 |
| **Past history** |  |  |
| Alcohol consumption | 2 | 3.9 |
| Tobacco consumption | 2 | 3.9 |
| Hypertension | 3 | 5.9 |
| Diabetes | 0 | 0 |
| Dyslipidemia | 0 | 0 |
| **Symptoms** |  |  |
| Asthenia | 4 | 7.8 |
| Fever | 11 | 21.6 |
| Anosmia | 8 | 15.7 |
| Aguesia | 6 | 11.8 |
| Headache | 5 | 9.8 |
| Articular pain | 4 | 7.8 |
| Thoracic pain | 2 | 3.9 |
| Dyspnea | 1 | 2 |
| Cough | 12 | 23.5 |
| Rhinorhea | 3 | 5.9 |

### Study endpoints

Table 2 shows the variations of ECG characteristics between D0, D3 and D7. Changes in QTc were not (429±27 ms at D0 and 396±26 ms at D7, p = 0.27). There were significant changes in the mean heart rate and the QRS duration between D0 and D7. No clinical arrhythmic events were observed during the treatment course. No death or sudden death were reported.

**Table 2:** Variations of electrocardiographic parameters at D0-D3, D0-D7, D3-D7.

| Variables | D0 | D3 | D7 | P value |  |  |
| --- | --- | --- | --- | --- | --- | --- |
|  |  |  |  | D0→D3 | D3→D7 | D0→D7 |
| **Mean heart rate±standard deviation (bpm)** | 75±13 | 78.8±14 | 70±13 | 0.72 | 0.23 | **0.02** |
| **Mean PR interval±standard deviation (msec)** | 170±24 | 175±30 | 183±38 | **0.002** | 0.94 | 0.25 |
| **Mean QRS duration ±standard deviation (msec)** | 95±10 | 93±12 | 102±17 | 0.1 | 0.73 | **0.004** |
| **Mean QTc duration ±standard deviation (msec)** | 429±27 | 434±29 | 396±26 | 0.61 | 0.38 | 0.27 |

bpm : beats per minute ; D : day; msec : millisecond.

## Discussion

Management of COVID-19 patients is a major challenge worldwide as no effective specific treatment is standard. The reported risk of ventricular arrhythmia has raised additional concerns in the use of Hydroxychloroquine and Azithromycin based protocols.^6,7^ While such protocols are extensively used in Africa since COVID-19 outbreak, little is known about the cardiac safety in this relatively young population with high likelihood of previous exposure to chloroquine or its derivatives in the fight against endemic malaria.

QT interval prolongation exposing to Torsarde de Pointes is a well-known side effect of hydroxychloroquine and Azithromycin. ^6,7,10–12^ The main hypothesis for QT prolongation in patients administered chloroquine or hydroxychloroquine is inhibition of the rapidly activating component of delayed rectifier potassium current (iKr). The arrhythmogenic mechanism of azithromycin is not very well established but it is hypothesized that azithromycin also acts through QT prolongation.^6^ In the COVID-19 context, some other pathological conditions linked to QT prolongation are: myocardial injury, renal failure, hepatic failure and electrolyte imbalance such as hypokalemia or hypomagnesemia. ^6,13^

Our findings in a young population with limited comorbidities and mild symptomatic COVID-19 do not support short term adverse effects on major ECG measurements, of Hydroxychloroquine at 400 mg daily when used in combination with Azithromycin. No clinically significant ECG changes nor arrhythmic events were observed during the treatment course. The fact that we did not find the significant modification of the QTc interval in the current study may be explained by the fact that all the patients had mild forms of COVID-19 and few co-morbid conditions. A risk score has been derived and validated by Tisdale *et al*, for prediction of drug-associated QT prolongation among cardiac-care-unit-hospitalized patients. A score ≥ 11 is highly predictive of QT prolongation during treatment, hence, risk of severe cardiac arrhythmia. Our population had a mean Tisdale score of 3 that predicted a low risk of cardiac arrhythmia. ^9^

We found a significant reduction in heart rate and an increase of QRS duration. Heart rate reduction could be the reflection of inflammation control by the immunomodulatory effect of hydroxychloroquine.^14^ Increase QRS duration even if it did not reach the set of definitions of QRS widening has also been reported in patients treated by hydroxychloroquine.^15^ These changes were not clinically significant in our cohort and could predict the safety of both hydroxychloroquine and azithromycin in non-severe forms of COVID-19.

## Study limitations

These results should be interpreted taking into account the limited sample size, the young age of the population with very few co-morbidities and the short term exposition to hydroxychloroquine and Azithromycin in the Cameroon national standard protocol to manage COVID-19 patients.

## Conclusions

We did not observe a significant increase in QTc-interval. Only mild and not clinically significant electrocardiographic abnormalities were observed in non-severe COVID-19 patients with low risk of arrhythmia assessed using the Tisdale score, treated with hydroxychloroquine and azithromycin in Cameroon.

## Data Availability

Data are available under request with prior approval by the Nationa Ethics Committee and the Scientific Advisory Board of Public Health Emergencies of Cameroon

## Acknowledgements

We acknowledge all the administrative staff of Public Health Delegation of Centre region, Yaounde Central Hospital, Yaounde Jamot Hospital, Djoungolo District Hospital, Health care providers of the COVID-19 treatment centres of these hospitals, research nurses, assistant secretaries and all patients who consented to participate in this study.

## Contributions

the authors all contributed equally.

## Ethical approval

The study was approved by the National Committee on Research Ethics for Human Health in cameroon before the conduct of the study (N°2020/05/1505/L/CNERSH/SP).

## Data sharing

No additional data available.

